# Healthcare utilisation among children in contact with social services in England: an interrupted time series using ECHILD

**DOI:** 10.64898/2026.01.09.26343784

**Authors:** Eliazar Luna, Lucy J Griffiths, Karen Broadhurst, Dougal Hargreaves, Jenny Woodman, Lisa Holmes, Kat Tranter, Grace A Bailey, Katie Harron

## Abstract

**Background:** Children in contact with children’s social care (CSC) services have high levels of hospital utilisation, but patterns before and after referral remain insufficiently understood.

**Objective:** To evaluate healthcare utilisation two years before and after CSC referral.

**Participants and setting:** Retrospective cohort using ECHILD linked health and social care data, including children with a first CSC referral between 2009 and 2018 in England.

**Methods:** We compared monthly planned and unplanned hospital contact rates for Children in Need, Children under Protection Plans, and Children Looked After with age-sex-matched cohorts. We used interrupted time series analysis to examine how healthcare utilisation changed following referral. We also explored reasons for hospital contacts.

**Results:** We analysed >12 million hospital contacts for 1,014,330 Children in Need, 204,240 Children under Protection Plan and 177,640 Children Looked After. Children Looked After had the highest average number of total contacts (11.8 per child over a 4 year period), followed by Children in Need (8.8) and Children under Protection Plans (8.4). All CSC groups had about twice the contacts of matched peers. Healthcare utilisation increased sharply prior to referral, with a peak around referral. After referral, planned care increased and unplanned care decreased, with pre-referral upward trends slowing or reversing. The most common reason for healthcare utilisation was mental health–related

**Conclusions:** At a population level, CSC referral marks a pivotal point in healthcare utilisation, with a shift from unplanned to planned care. This may reflect more structured engagement with health services and coordinated support for children and families.

## 1. Introduction

Children’s social care services (CSC) provide support to children, young people and families who experience adverse circumstances. In England, local authorities have statutory responsibilities regarding the care of these children (Department for Education, 2021). Children involved with CSC receive different types of support depending on their circumstances, including support as Children in Need (CiN), protection through Child Protection Plans (CPP), or care as Children Looked After (CLA). We use statutory CSC terms to align with administrative data and legal definitions, while recognising this language may feel impersonal or stigmatising, and not reflect how children and young people see their own lives (Marcolino et al., 2018). The Department for Education (DfE) reported a total of 399,460 Children in Need, 49,900 Children on a Child Protection Plan and 83,630 Children Looked After in 2024 (Department for Education, 2024a, 2024b). Before turning 18, 25.9%, 6.9% and 3.3% of all children in England are supported at least once as Children in Need, spent time on a Protection Plan or are looked after by their local authority, respectively (M. Jay et al., 2024; Mc Grath-Lone, Dearden, et al., 2016). Across all groups, abuse or neglect is the predominant category for CSC involvement (Department for Education, 2024a, 2024b). Such experiences place children at increased risk of poor outcomes, including for health.

Evidence has consistently shown that children in contact with CSC experience worse health outcomes, and have higher levels of healthcare utilisation than their peers (Allik et al., 2021, 2024; Baldwin et al., 2024; Fleming et al., 2021; Lowthian et al., 2024; McCue Horwitz et al., 2012; McKenna et al., 2023; Rubin et al., 2004; Soraghan & Porter, 2024; Szilagyi et al., 2015). Population-level studies in Scotland have shown a higher risk of hospitalisation, self-harm and injuries for Children Looked After compared to their peers (Fleming et al., 2021), as well as higher rates of use of outpatient services, mental health services and A&E visits (Allik et al., 2021). Population-level cohort studies in Wales and Northern Ireland have highlighted that the increased healthcare utilisation compared to general population extends to all CSC groups and not just Children Looked After (Lowthian et al., 2024; McKenna et al., 2023). Observational studies from Australia and the US have also reported greater emergency healthcare utilisation for children in out of home care compared to peers (Baldwin et al., 2024; Rubin et al., 2004). Despite higher healthcare utilisation, involvement with CSC may help to support health management and promote a shift from reactive to planned care (Ross et al., 2004). Some evidence has found CSC involvement to be associated with increased use of inpatient and subspecialty services, but not emergency services, compared with matched controls (Bennett et al., 2020). Population-wide studies have reported an increased risk of hospitalisations before and after care episodes but not during (Allik et al., 2024). This suggests distinct patterns of healthcare utilisation during children’s social care trajectories. The role of CSC in shifting unplanned to planned care is important because unplanned care is linked to poorer outcomes (Coughlan et al., 2022), and because a small number of children drive a disproportionate share of healthcare utilisation, most of it unplanned (Beaney et al., 2021). However, outcomes in CSC are not only about reducing unplanned care but also about safety, stability, health and developmental progress (Forrester, 2017; La Valle et al., 2019). Children referred to CSC often present with overlapping adversities, which shape their needs and the meaning of improvement (Hood et al., 2024). In this context, more planned care may reflect not just improved health management but also more coordinated, valued support for children and families.

Understanding how healthcare patterns emerge and evolve in relation to CSC involvement is increasingly recognised as a priority, yet key gaps remain. Research disproportionately focuses on children in out of home care, with children who remain with their families with CSC support being underrepresented. Existing studies predominantly examine mental over physical health (Bailey et al., 2025). Most research focuses on healthcare utilisation during or after CSC involvement, with less focus on timing and pre-referral patterns (Allik et al., 2024). As a result, it remains unclear whether high healthcare utilisation reflects pre-existing need or changes following referral and CSC intervention. Stakeholders and researchers emphasise the importance of data-driven longitudinal, cross-sectoral analyses that offer a more comprehensive view of children’s journeys through services and their outcomes, as well as generate insights to evaluate service effectiveness. (Griffiths et al., 2025; Tranter et al., 2025).

In this study, we address these priorities by using linked administrative data to describe and model healthcare utilisation patterns in the two years before and after first CSC involvement and comparing them with matched peers.

## 2. Methods

### 2.1 Study Design

This is a retrospective cohort design using an interrupted time series (ITS). The ITS design is well-suited for describing changes associated with interventions occurring at a distinct point in time and enables separate estimation of changes in outcome level and trend (Lopez Bernal et al., 2016).

We followed the RECORD (REporting of studies Conducted using Observational Routinely-collected health Data) (Benchimol et al., 2015) guidelines to ensure comprehensive reporting of methods, data handling, and analysis.

### 2.2 Data source

We used the Education and Child Health Insights from Linked Data (ECHILD), which links together administrative records from hospitals, schools and social care services for children, for ∼20 million children in England (Mc Grath-Lone et al., 2022). Within ECHILD, we used health data captured in Hospital Episode Statistics (HES) and CSC data (Herbert et al., 2017; M. A. Jay et al., 2019). The study population consisted of children involved with CSC, recorded using defined administrative codes in the Children in Need and Children Looked After Census data in ECHILD (Emmott et al., 2019; Mc Grath-Lone, Harron, et al., 2016), who linked to HES.

### 2.3 Participants

Eligible children were Children in Need, Children on a Child Protection Plan and Children Looked After, first in contact with CSC in England between April 2009 and March 2018. Children were included if referred to CSC in the study period, aged ≥2 and <18 years old when their CSC episode started. Children were considered Children in Need if they had been assessed and found in need for the first time during this period without having a Children on a Protection Plan or a Children Looked After episode within two years of Children in Need episode start date.

Children on a Protection Plan must have been placed on a protection plan for the first time during this period and not have had a Children Looked After episode within two years of Children on a Protection Plan episode start date. Children were considered Children Looked After if they had their first recorded placement during this period.

We excluded those with missing referral date, those aged <2 or ≥18 years old, those with missing ethnicity, deprivation or age (3% missingness in sociodemographic variables), and those first referred outside the study period. Where there were duplicate records for the same individual, the earliest observation was used. We also excluded those who did not have an anonymised pupil matching reference number or a Token Person ID (required for linkage between NPD and HES). Approximately 5% of eligible children were not linked to a HES record (**Supplementary Material A and B).**

### 2.4 Outcomes

The primary outcome was the monthly number of hospital contacts for A&E, unplanned admissions, planned admissions, and outpatients, grouped as planned (outpatients, planned admissions) and unplanned care (A&E, unplanned admissions).

### 2.5 Statistical methods

We described the sociodemographic characteristics of the sample. We then examined linkage rates between CSC and HES data by year of referral.

The intervention point in this study was defined as the date of CSC referral, with each individual’s time series centred around this date (month 0). We used data for each child for a period of 48 months — 24 months before and 24 months after referral. We estimated the total number, mean, median, and monthly rate (per 1000 children) of hospital contacts in the two years before and after referral to CSC. We also used outpatient data to describe the rate and distribution of outpatient specialties attendances, and inpatient data to describe diagnoses of chronic conditions (Hardelid et al., 2014). Analyses were performed separately for each CSC group, and compared with 1:1 age-sex matched cohorts of children with no recorded CSC involvement, with each matched child assigned the same time 0 as their CSC peer.

For the ITS, we excluded the period between months -2 to 2 due to a distinctive spike in healthcare utilisation around the time of referral (this period was retained for descriptive analyses). We fitted negative binomial regression models to estimate changes in the level and slope of hospital contact rates before versus after referral, (Hilbe, 2011).

In adjusted analyses, we accounted for age at referral (2–4 years, 5–11 years, and 12–17 years), sex (male or female), ethnicity (White, Black, Asian, Mixed, and Any Other ethnic group; according to modal ethnicity across available CSC episodes) and area-level deprivation measured using the 2019 Index of Multiple Deprivation (IMD) of the local authority, categorised into deciles (Ministry of Housing, Communities & Local Government, 2019).

All analyses were performed in R version 4.3. The full analysis code and specifications for data cleaning and model fitting are available in the ECHILD GitHub repository (GitHub, n.d.).

## 3. Results

We included 1,014,330 Children in Need, 204,240 Children on a Child Protection Plan and 177,640 Children Looked After involved with CSC for their first time during the study period who linked with a health record. Between 94-95% of children involved with CSC linked to a hospital record; there were no substantive differences over time or in the sociodemographic characteristics of the sample before and after linkage (**Supplementary B, C)**. Children on Protection Plans were on average younger (mean age 8.4 years) than Children in Need (9.7 years) and Children Looked After (10.9 years; **Table 1**). The majority of children of CSC groups had abuse or neglect as their primary need codes. Our sample included about twice as many children from the most deprived decile compared to the least deprived.

**Table 1.** Characteristics of Children in Need, Children on a Child Protection Plan and Children Looked After with a first referral to CSC between April 2009 and March 2018, aged ≥2 and <18 years

| Variable | Category | CiN (N = 1,014,330) |  | CPP (N = 204,240) |  | CLA* (N = 177,640) |  |
| --- | --- | --- | --- | --- | --- | --- | --- |
|  |  | N | % | N | % | N | % |
| Year of referral | 2009-2010 | 171,830 | 16.9 | 2,730 | 1.3 | 34,060 | 19 |
|  | 2010-2011 | 123,600 | 12.2 | 7,860 | 3.8 | 21,400 | 12 |
|  | 2011-2012 | 118,730 | 11.7 | 21,110 | 10.3 | 18,550 | 10 |
|  | 2012-2013 | 118,300 | 11.7 | 25,510 | 12.5 | 17,460 | 10 |
|  | 2013-2014 | 114,610 | 11.3 | 28,710 | 14.1 | 17,770 | 10 |
|  | 2014-2015 | 95,530 | 9.4 | 28,480 | 13.9 | 17,450 | 10 |
|  | 2015-2016 | 90,700 | 8.9 | 28,290 | 13.9 | 16,840 | 9 |
|  | 2016-2017 | 89,150 | 8.8 | 29,600 | 14.5 | 17,320 | 10 |
|  | 2017-2018 | 91,890 | 9.1 | 31,940 | 15.6 | 16,790 | 9 |
| Continuous age | Mean years (SD) | 9.7 | (4.4) | 8.4 | (4.0) | 10.9 | (4.7) |
| Age group | 2 - 4 years old | 182,590 | 18.0 | 51,960 | 25.4 | 28,560 | 16 |
|  | 5 – 11 years old | 485,060 | 47.8 | 105,920 | 51.9 | 63,920 | 36 |
|  | 12 -17 years old | 346,680 | 34.2 | 46,360 | 22.7 | 85,150 | 48 |
| Ethnicity | Any other ethnic group | 20,130 | 2.0 | 2,940 | 1.4 | 3,900 | 2 |
|  | Asian | 83,580 | 8.2 | 14,500 | 7.1 | 10,020 | 6 |
|  | Black | 73,840 | 7.3 | 11,230 | 5.5 | 15,090 | 8 |
|  | Mixed | 60,320 | 5.9 | 16,250 | 8 | 15,010 | 8 |
|  | White | 776,460 | 76.5 | 159,320 | 78 | 133,620 | 75 |
| Sex | Male | 520,130 | 51.3 | 104,130 | 51 | 96,320 | 54 |
|  | Female | 494,210 | 48.7 | 100,100 | 49 | 81,320 | 46 |
| Primary need code | Abuse or neglect | 519,510 | 51.2 | 126,480 | 61.9 | 98,300 | 55 |
|  | Child's disability/illness | 60,990 | 6.0 | 3,550 | 1.7 | 10,640 | 6 |
|  | Parental disability/illness | 23,280 | 2.3 | 5,550 | 2.7 | 5,680 | 3 |
|  | Family in acute stress | 106,540 | 10.5 | 17,130 | 8.4 | 18,410 | 10 |
|  | Family dysfunction | 188,440 | 18.6 | 39,860 | 19.5 | 29,710 | 17 |
|  | Socially unacceptable behaviour | 27,690 | 2.7 | 3,500 | 1.7 | 6,470 | 4 |
|  | Low income/ Absent parenting | 13,220 | 1.3 | 1,290 | 0.6 | 8,440 | 5 |
|  | Cases other than CiN | 13,460 | 1.3 | 1,270 | 0.6 | - | - |
|  | NA | 61,210 | 6.0 | 5,600 | 2.7 | - | - |
| **Source** | Individual | 54,120 | 5.3 | 12,540 | 6.1 | - | - |
|  | Schools | 126,850 | 12.5 | 37,740 | 18.5 | - | - |
|  | Health services | 63,640 | 6.3 | 19,800 | 9.7 | - | - |
|  | Housing | 6,070 | 0.6 | 1,860 | 0.9 | - | - |
|  | LA services | 60,090 | 5.9 | 27,420 | 13.4 | - | - |
|  | Police | 134,660 | 13.3 | 46,820 | 22.9 | - | - |
|  | Other legal agency | 14,220 | 1.4 | 4,480 | 2.2 | - | - |
|  | Other | 32,420 | 3.2 | 11,100 | 5.4 | - | - |
|  | Anonymous | 8,960 | 0.9 | 3,900 | 1.9 | - | - |
|  | NA | 513,300 | 50.6 | 38,580 | 18.9 | - | - |
| **IMD Decile** | 1 Most deprived | 127,410 | 12.6 | 26,460 | 13.0 | 25,080 | 14 |
|  | 2 | 83,490 | 8.2 | 18,260 | 8.9 | 15,790 | 9 |
|  | 3 | 81,890 | 8.1 | 15,180 | 7.4 | 15,580 | 9 |
|  | 4 | 96,770 | 9.5 | 20,220 | 9.9 | 18,400 | 10 |
|  | 5 | 92,400 | 9.1 | 19,460 | 9.5 | 17,550 | 10 |
|  | 6 | 106,390 | 10.5 | 17,240 | 8.4 | 16,780 | 9 |
|  | 7 | 149,930 | 14.8 | 30,420 | 14.9 | 22,600 | 13 |
|  | 8 | 94,780 | 9.3 | 18,560 | 9.1 | 17,540 | 10 |
|  | 9 | 109,620 | 10.8 | 23,220 | 11.4 | 18,230 | 10 |
|  | 10 Least deprived | 71,670 | 7.1 | 15,230 | 7.5 | 10,090 | 6 |

*\*Children Looked After percentages have been rounded to 0 d.p in compliance with DfE statistical disclosure policy*

In the two years prior to referral, there were an average of 4.09 (Children in Need), 3.71 (Children on a Child Protection Plan) and 5.21 (Children Looked After) hospital contacts per child (median = 1, 1 and 2, respectively) (**Table 2**). Children Looked After had a higher mean number of hospital contacts across all health services. Outpatient appointments accounted for the majority of contacts. A&E was the next most frequently used service, particularly for Children Looked After. Unplanned admissions were more frequent than planned admissions for Children Looked After. For Children in Need, planned and unplanned admissions were very similar, while among Children under Protection Plans, unplanned admissions exceeded planned admissions. Overall, children with no CSC involvement had around half of the hospital contacts of those involved with CSC, but this difference increased at the time of referral, particularly for unplanned admissions, where children looked after had nearly seven times the rate of their matched peers (**Supplementary Material D,** Table S5).

**Table 2.** Average monthly rate, mean and median hospital contacts by time period, CSC groups and health service

| Type of care | Health service | CSC group | Average monthly rate of hospital contacts per 1,000 children 2 years pre CSC referral |  | Average monthly rate of hospital contacts per 1,000 children 2 years post CSC referral |  | Mean (median) number of contacts per child 2 years pre CSC referral |  | Mean (median) number of contacts per child 2 years post CSC referral |  |
| --- | --- | --- | --- | --- | --- | --- | --- | --- | --- | --- |
|  |  |  | Children involved with CSC | Matched peers | Children involved with CSC | Matched peers | Children involved with CSC | Matched peers | Children involved with CSC | Matched peers |
| Unplanned care | A&E Attendances | CiN | 37.81 | 23.79 | 39.68 | 23.32 | 0.91 (0) | 0.57 (0) | 0.95 (0) | 0.56 (0) |
|  |  | CPP | 40.12 | 26.76 | 40.67 | 24.73 | 0.96 (0) | 0.64 (0) | 0.98 (0) | 0.59 (0) |
|  |  | CLA | 47.08 | 24.08 | 59.30 | 23.70 | 1.13 (0) | 0.58 (0) | 1.43 (0) | 0.57 (0) |
|  | Unplanned admissions | CiN | 8.35 | 3.80 | 7.96 | 3.08 | 0.20 (0) | 0.09 (0) | 0.19 (0) | 0.07 (0) |
|  |  | CPP | 7.95 | 4.51 | 7.77 | 3.34 | 0.19 (0) | 0.11 (0) | 0.19 (0) | 0.08 (0) |
|  |  | CLA | 12.96 | 3.76 | 12.78 | 3.14 | 0.31 (0) | 0.09 (0) | 0.31 (0) | 0.08 (0) |
| Planned care | Planned admissions | CiN | 7.24 | 3.40 | 9.08 | 3.54 | 0.17 (0) | 0.08 (0) | 0.22 (0) | 0.09 (0) |
|  |  | CPP | 4.89 | 3.56 | 6.33 | 3.57 | 0.12 (0) | 0.09 (0) | 0.15 (0) | 0.09 (0) |
|  |  | CLA | 9.15 | 3.29 | 10.04 | 3.62 | 0.22 (0) | 0.08 (0) | 0.24 (0) | 0.09 (0) |
|  | Outpatient appointments | CiN | 116.94 | 62.99 | 141.09 | 68.80 | 2.81 (0) | 1.51 (0) | 3.39 (0) | 1.66 (0) |
|  |  | CPP | 101.65 | 66.16 | 140.64 | 71.27 | 2.44 (0) | 1.59 (0) | 3.38 (1) | 1.72 (0) |
|  |  | CLA | 147.80 | 64.58 | 193.76 | 71.30 | 3.55 (0) | 1.55 (0) | 4.66 (2) | 1.71 (0) |
| Combined hospital contacts |  | CiN | 170.34 | 93.98 | 197.81 | 98.74 | 4.09 (1) | 2.26 (0) | 4.75 (1) | 2.37 (0) |
|  |  | CPP | 154.61 | 100.99 | 195.41 | 102.91 | 3.71 (1) | 2.43 (1) | 4.70 (2) | 2.48 (1) |
|  |  | CLA | 216.99 | 95.71 | 275.88 | 101.76 | 5.21 (2) | 2.30 (1) | 6.63 (3) | 2.45 (0) |

Two years after referral, the average number of contacts per child increased to 4.75 (Children in Need), 4.70 (Children on a Protection Plan) and 6.63 (Children Looked After). For all CSC groups, rates of A&E attendances, planned admissions and outpatient appointments were higher in the two years after CSC referral compared to the two years before (**Table 2**). Conversely, the rate of unplanned admissions was lower after CSC referral. There were particularly large increases in average monthly rates of planned admissions for Children in Need (25.4% increase), and outpatient appointments for Children on a Protection Plan and Children Looked After (38.4% and 31.1% increase). Unplanned admissions decreased post referral by 4.7%, 2.3%, and 1.4% for Children in Need, Children on a Protection Plan and Children Looked After, respectively (**Table 2**). Details on the number of contacts by group and period are described in **Supplementary Material D**.

Before CSC referral, monthly hospital contact rates were increasing over time in most CSC groups and services (**Figure 1, Supplementary material E,** Tables S12 to S17). The steepest increase was observed for unplanned admissions among Children Looked After, rising by 1.20% per month (26.8% yearly). The gradual pre-referral change was interrupted by a substantial peak around CSC referral (grey area), which declined within a few months before stabilising (**Figures 1** and **2**). The largest peak in hospital contacts was seen in Children Looked After. Planned care tended to peak at or just after CSC involvement; unplanned care peaked just before or at CSC involvement and quickly decreased right after, returning to pre-referral rates. Hospital contact rates in matched peers were relatively stable and the gap with their CSC counterparts was smallest for Children on a Protection Plan, larger for Children in Need and largest for Children Looked After (**Figures 1** and **2**).

**Figure 1:**
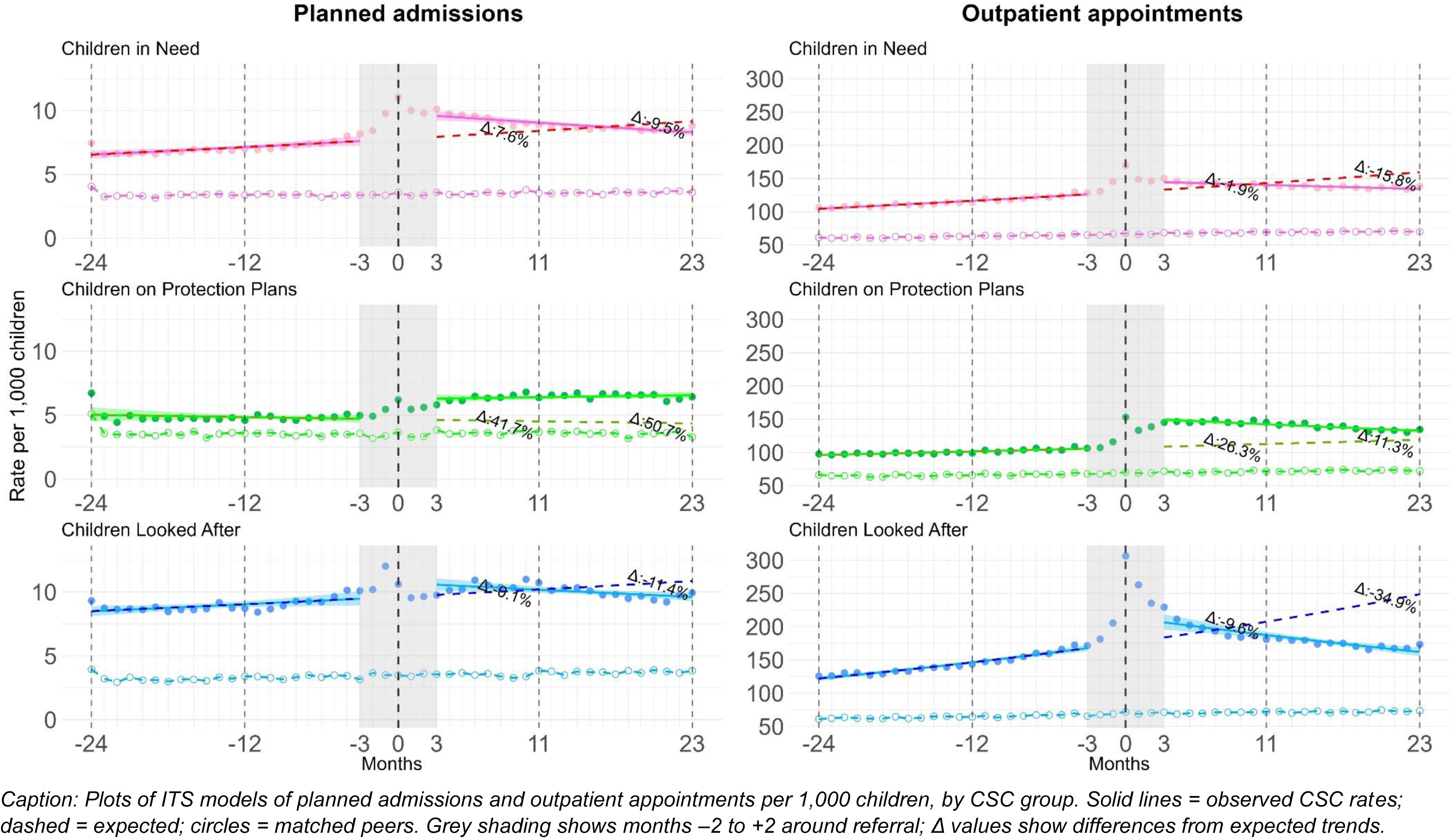
**Observed and expected rates of planned hospital contacts by CSC groups and matched peers** Caption: Plots of ITS models of planned admissions and outpatient appointments per 1,000 children, by CSC group. Solid lines = observed CSC rates; dashed = expected; circles = matched peers. Grey shading shows months –2 to +2 around referral; Δ values show differences from expected trends.

**Figure 2:**
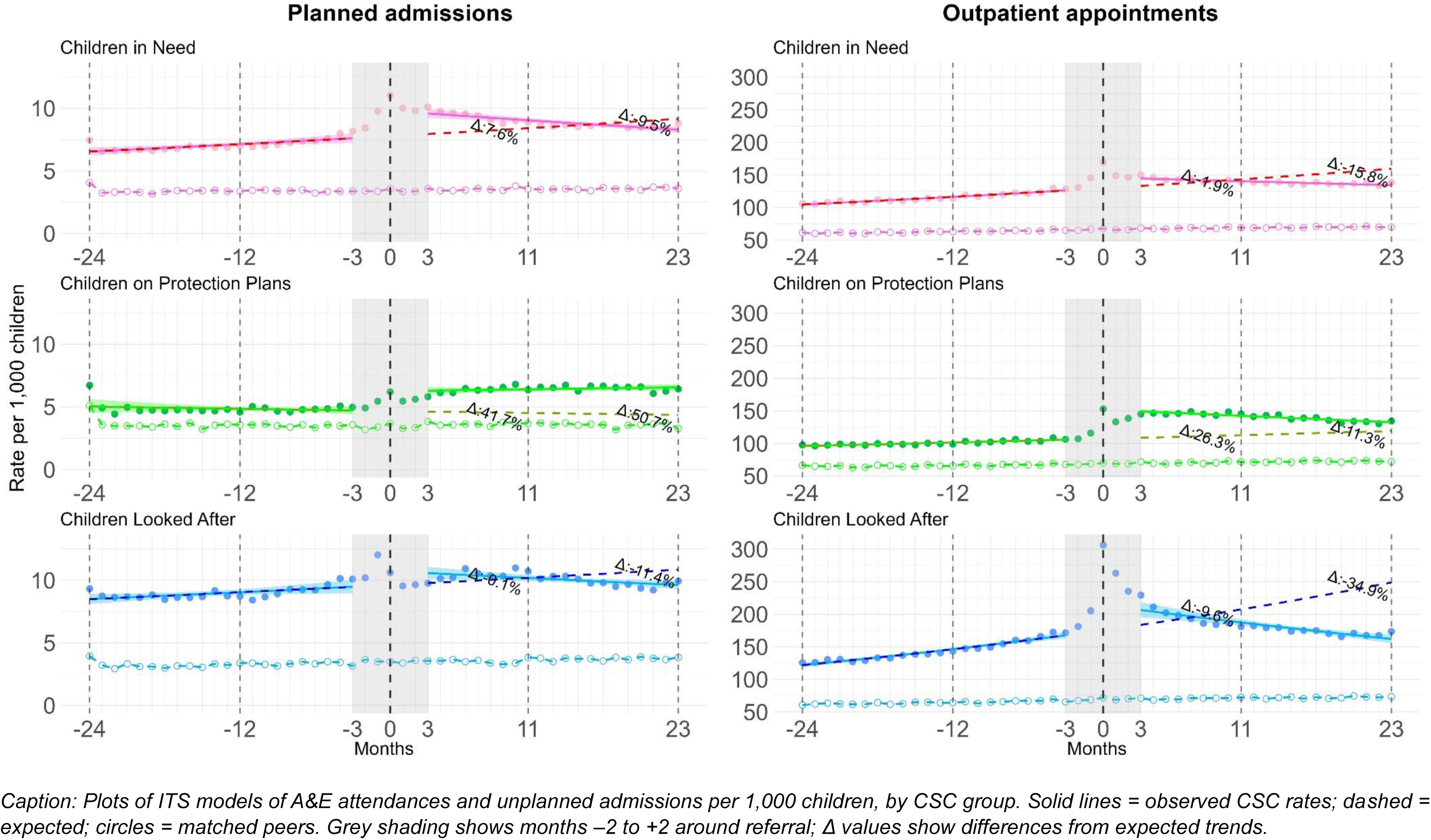
Observed and expected rates of unplanned hospital contacts by CSC groups and matched peers *Caption: Plots of ITS models of A&E attendances and unplanned admissions per 1,000 children, by CSC group. Solid lines = observed CSC rates; dashed = expected; circles = matched peers. Grey shading shows months –2 to +2 around referral; Δ values show differences from expected trends*.

There was a level change in the number of hospital contacts following CSC referral, particularly for planned care. For example, for Children on Protection Plans, there was a 41.8% (95% CI 1.37 – 1.46) increase in the monthly rate of outpatient appointments after CSC referral. After CSC referral, the rate of increase in hospital contacts over time generally slowed (**Supplementary Material E,** Tables S6 to S17). This is illustrated in **Figure 2**, where the slope in unplanned care is flatter following referral, especially for Children in Need and Children Looked After. Hospital contact rates varied significantly by age, sex, deprivation, and ethnicity (**Supplementary Material E** Tables S6 to S17).

The observed versus expected rates of hospital contacts (had pre referral trends continued), for Children in Need and Children Looked After diverged over time. Two years post-referral, the observed rate of unplanned admissions for Children Looked After was 54.3% lower than the predicted rate without CSC involvement (**Figure 2**). For Children on Protection Plans, unplanned care was initially lower in the model of no CSC involvement, but the gap narrowed over time. For planned care, Children in Need and Children Looked After also had similar patterns: the model based on pre referral trends shows a continued upward trend over time. The full model showed rates steadily declining after referral. A year post referral, the monthly rates in both models were similar, and after two years, the predicted rates were lower in the model based on pre referral trends.

Excluding Paediatrics, the most frequently used outpatient specialty among CSC groups was Child and Adolescent Psychiatry, followed by Trauma & Orthopaedics and Ophthalmology (**Figure 3**). This pattern was not mirrored among matched peers. Outpatient service use increased across most specialities after referral among CSC groups, except for Paediatric Neurology and Ear, Nose, and Throat. The largest increases were seen in Child and Adolescent Psychiatry and Learning Disability services, which also showed the greatest differences compared with matched peers. Overall, CSC groups had higher rates of outpatient use across all specialties except for Orthodontics. The burden of chronic conditions, measured as the rate of diagnoses, was consistently higher in CSC groups than in their matched peers across all categories (**Figure 4**). Similar to outpatient services, the most common diagnosed conditions were related to mental health. Following referral, diagnosis rates declined for all categories except mental health/behavioural conditions, a pattern also observed in matched peers. The largest changes among CSC groups occurred in neurological and mental health/behavioural conditions, which were also the categories with the largest gaps in diagnosis rates between CSC groups and matched peers.

**Figure 3.**
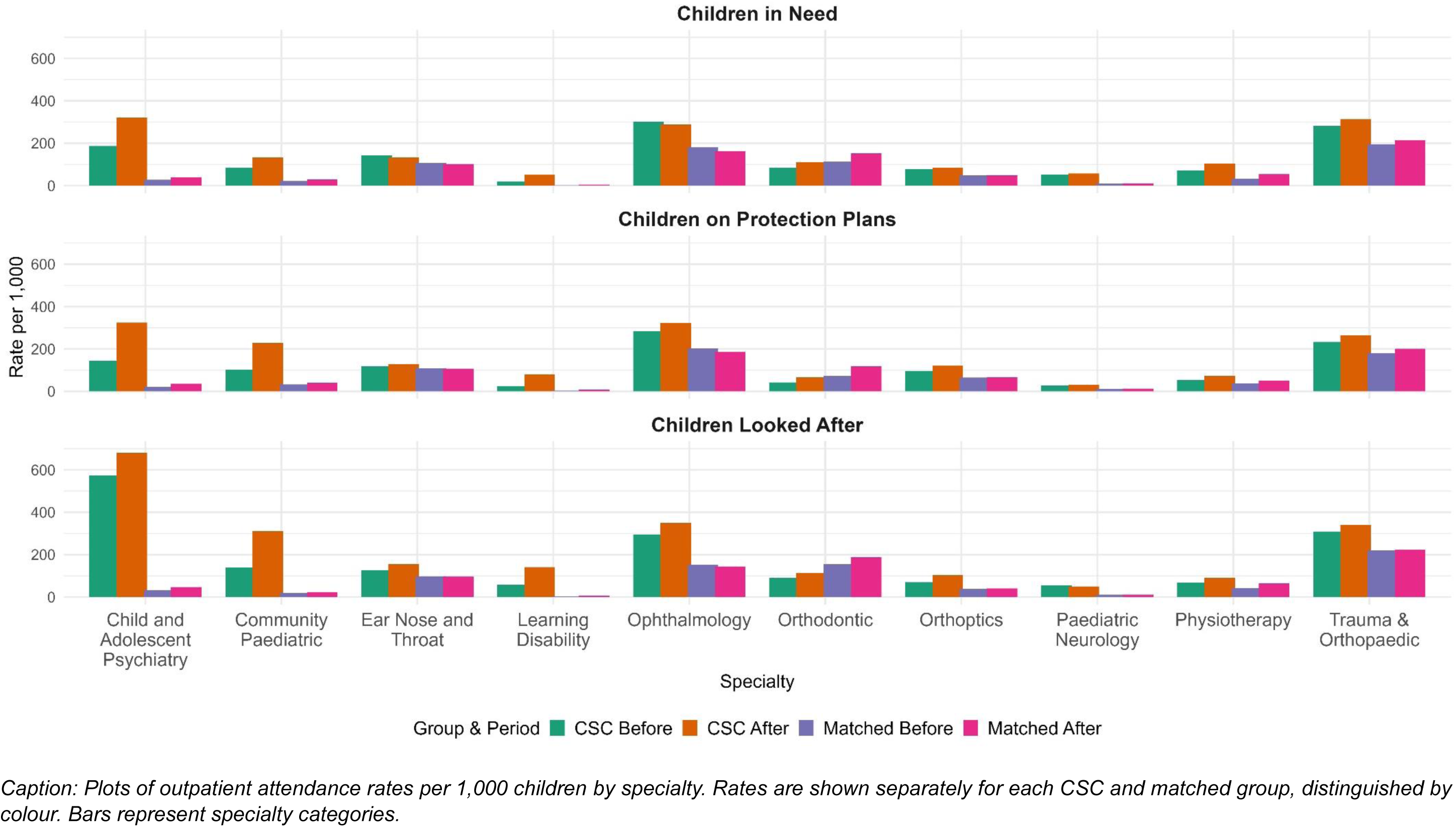
Outpatient attendance rates in top 10 specialties (excluding Paediatrics): before and after CSC referral compared to matched peers ***Caption: Plots of outpatient attendance rates per 1,000 children by specialty. Rates are shown separately for each CSC and matched group, distinguished by colour. Bars represent specialty categories*.**

**Figure 4.**
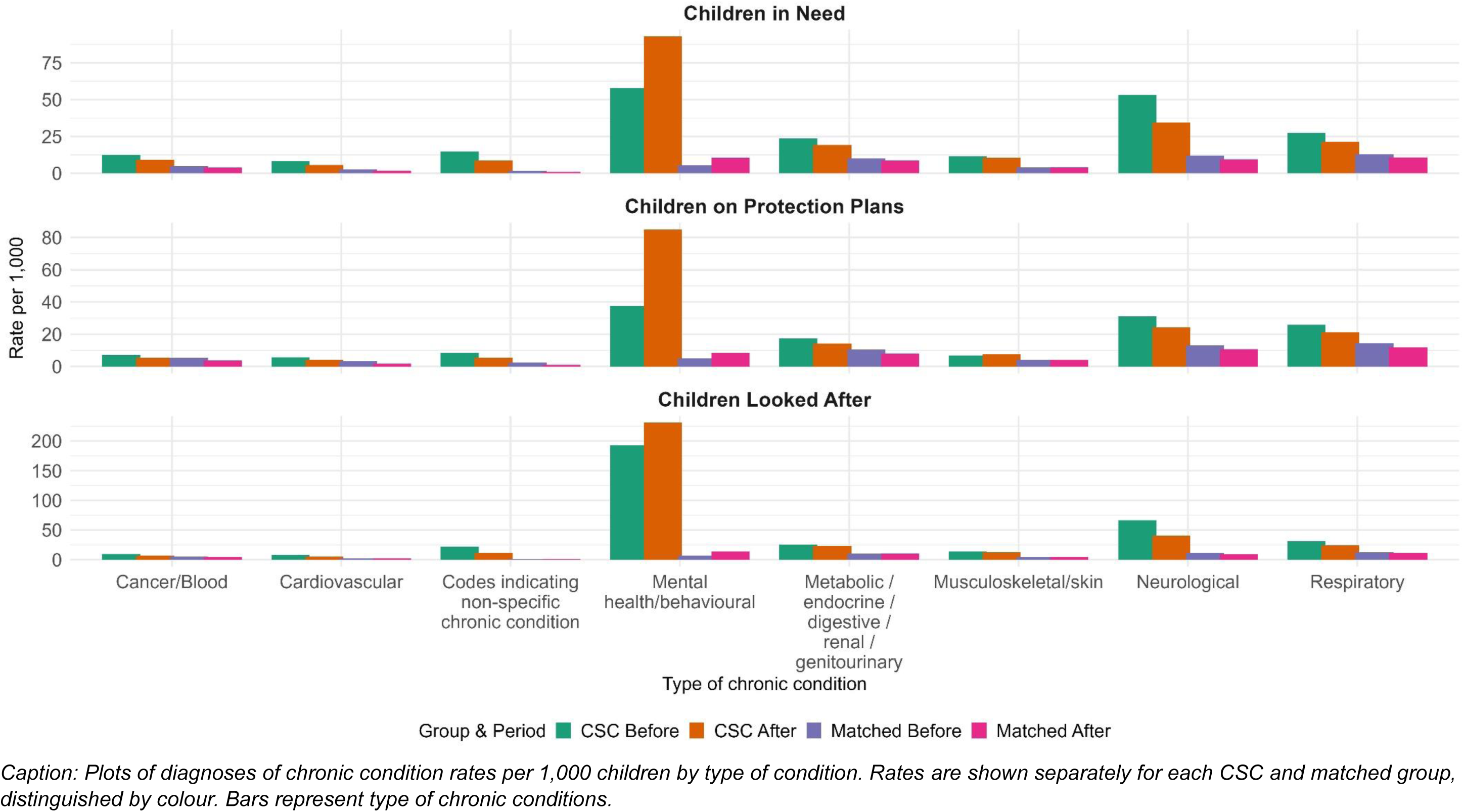
Rates of diagnoses of chronic conditions by type: before and after CSC referral compared to matched peers *Caption: Plots of diagnoses of chronic condition rates per 1,000 children by type of condition. Rates are shown separately for each CSC and matched group, distinguished by colour. Bars represent type of chronic conditions*.

## 4. Discussion

In our study of >1 million children involved with CSC in England, we found that hospital contacts consistently peak around the time of CSC involvement, highlighting this as a period of high health need and/or more service engagement. We also found that trends in hospital contacts changed substantially post referral, with an immediate increase in planned care, and gradual decrease in the rate of unplanned care. While this pattern may reflect the more structured engagement with healthcare services that can follow CSC involvement, it could also reflect broader health-system responses triggered by the identification of welfare concerns, rather than the direct effects of CSC itself. The trend in hospital contact rates shifted at referral: the pre-referral upward trend was either slowed, flattened, or reversed. Across the examined period, hospital contact rates were higher after CSC involvement for all services except unplanned admissions. For all groups, children with CSC involvement had roughly double the level of healthcare utilisation compared to their matched peers, with mental health–related care being the predominant reason. Children Looked After showed the highest overall healthcare utilisation, with patterns similar to Children in Need but on a greater scale

This study has a number of limitations. First, it relies on observational data not originally collected for research purposes, affected by misclassification, unmeasured confounding, and changes in data quality over time. Also, data linkage between health and social care data for those under five is limited to those who remain involved with CSC during their school years (Mc Grath-Lone, 2017). While we adjusted for key sociodemographic factors, we lacked data on other important elements such as parental mental health and household-level deprivation. Our ITS models assume that, in the absence of CSC involvement, trends in hospital contacts would have continued the same, which may not hold. However, the analyses still provide strong evidence that CSC involvement substantially changed the trajectory of healthcare utilisation. We were also unable to account for the type of actual support received by children, as local authorities are not required to submit this. Our results provide a national picture of healthcare utilisation, but we acknowledge that individuals’ experiences are diverse, and CSC referral may have no impact on healthcare utilisation for some. Finally, our analysis did not include primary care. We recognise its importance, especially in early intervention, and future research should include other health services to get a more comprehensive view of healthcare utilisation.

On the other hand, our study also has strengths. The ITS allows us to estimate both immediate and gradual changes, helping to separate short-term spikes around referral from longer-term trends. Our samples provided sufficient statistical power and national representativeness. The creation of matched cohorts helped to decrease the likelihood of findings being explained by the effect of age on healthcare utilisation. Our analysis extends the evidence beyond children in out of home care to Children in Need and Children on a Child Protection Plan. In line with stakeholder priorities (Tranter et al., 2025), we moved beyond descriptive work to provide a longitudinal view of healthcare utilisation in relation to CSC referral. Our estimates remained consistent in adjusted analyses, suggesting that changes were not driven by sociodemographic factors. The generalisability of our findings is supported by the use of population-wide data from ECHILD, which includes most children in England and high linkage rates across health and social care data. This supports the applicability of the results to children involved with CSC across the country.

The elevated hospital contact rates observed around the time of referral are consistent with prior studies of children involved with CSC (Allik et al., 2021, 2024; Baldwin et al., 2024; Fleming et al., 2021; Lowthian et al., 2024; McCue Horwitz et al., 2012; McKenna et al., 2023; Rubin et al., 2004; Soraghan & Porter, 2024; Szilagyi et al., 2015). Some evidence shows that A&E attendances cluster after placement changes among Children Looked After (Rubin et al., 2004), suggesting that transitions in care can be disruptive. Our findings build on this by showing that transitions into different levels of support are also a critical window of healthcare utilisation, affecting not only children in out of home care but also those who remain with their families with CSC support. Our findings also show large differences in healthcare utilisation between children with and without CSC involvement. This gap has been described before in other UK nations, including Scotland, Wales and Northern Ireland, but mostly for Children Looked After (Fleming et al., 2021; Lowthian et al., 2024; McKenna et al., 2023). We show that elevated healthcare utilisation is high across all types of CSC involvement compared to peers, with the widest gaps for older children around the time of CSC referral. The increase in healthcare utilisation for Children in Need and Children on a Child Protection Plan calls attention to the healthcare needs of children outside the looked-after system, a group often still overlooked in both policy and research (Griffiths et al., 2025). The gap between the mean and median number of hospital contacts indicates a skewed distribution, consistent with evidence that healthcare utilisation is concentrated within small groups (Beaney et al., 2021). It is also important to emphasise that healthcare utilisation is an imperfect proxy for health needs, and a significant treatment gap exists. For example, about 30% of children and young people with a mental health condition access treatment (NHS, 2019). Therefore, an increase in healthcare utilisation may not indicate worse health, but rather it could mean better identification and proactive management of health needs, especially if these contacts are for planned care. Studies show clear contrasts between planned and unplanned healthcare use among children in care (Blackburn et al., 2025), and unplanned care is often associated with poorer health outcomes (Coughlan et al., 2022). Evidence also indicates that stronger continuity and integration between social and healthcare services can reduce unplanned care and improve outcomes (Curry & Ham, 2012; Hoffmann et al., 2023). Although NICE guidelines emphasise the importance of coordinated and continuous healthcare for children in care (National Institute for Health and Care Excellence, 2021), these standards are not always achieved in practice.

Prior research using administrative data has shown that children with CSC involvement are more likely to use mental health services (Fleming et al., 2021; McKenna et al., 2023), while there is also survey data evidence suggesting the opposite (Stein et al., 2016). Our results support the former interpretation: we observed an increase in the rate of mental health outpatient contacts and mental/behavioural chronic diagnoses after referral. Mental health was the main reason for healthcare utilisation among children with CSC involvement, a pattern not observed in their matched peers. We expanded the current evidence by exploring this difference for all types of CSC support and examining differences not just after, but before CSC involvement as well.

The shift from unplanned to planned care suggests that once children are involved with CSC, their healthcare needs may begin to be addressed in a more structured and proactive way. This aligns with other research showing reduced A&E visits and increased primary care use among children with social worker support, though their sample of a hundred children limits generalisability (Ross et al., 2004). For Children Looked After, this shift is somewhat expected, as there are statutory Review Health Assessments required at entry to care and then every 6 months (Department for Education, 2022; UK Statutory Instruments, 2010). However, the evidence on how effectively these assessments identify and address health needs remains limited (Herlitz et al., 2024; Hill & Watkins, 2003). Notably, we also observe this shift towards planned care among those with less formal support, such as Children in Need.

Retrospective cohorts studies have also reported higher inpatient and subspecialty use without increased emergency care in out of home care children (Bennett et al., 2020). Our study builds on this by using a large sample and including all CSC groups, suggesting a broader redirection toward planned healthcare, not just for Children Looked After but for all types of CSC support. This shift may also have economic implications since structured access to planned services can prevent the escalation of health issues that might otherwise result in avoidable A&E attendances or unplanned admissions (Keeble & Kossarova, 2017; Purdy, 2010). Improved coordination of social and health care services can contribute not only to improved outcomes but also towards a more sustainable health system (OECD, 2015).

Evidence suggests that proactive, early engagement with families can mitigate crises that would otherwise lead to intensive interventions (Chowdry & Oppenheim, 2018). Our study suggests that, in the long run, healthcare utilisation is generally lower than it would have been had pre CSC referral trends continued. We recognise the limitation of this claim, therefore, we prefer to highlight that, at the population level, CSC involvement changed the healthcare trajectory of children. This may indicate a protective effect of CSC involvement, with structured support enabling better health management. However, this could also reflect health services responding to increased awareness of social welfare concerns, rather than a direct protective effect of CSC. Despite the reasons for the shift, interpreting it requires caution. Patterns of service use often reflect differences in local provision and thresholds, rather than levels of need (Local Government Association & Newton, 2018). Also, while a rise in planned care may help reduce reliance on unplanned care, this would only lead to fiscal benefit if support is timely, sustained, and appropriately matched to children’s needs (Chowdry & Oppenheim, 2018; Feinstein et al., 2017).

The underlying drivers of the shift from unplanned to planned care following referral in these groups remain unclear. One possibility is that reductions in unplanned care could reflect greater stability in children’s environments (Rubin et al., 2004) or improvements in determinants of health following referral (Sandel et al., 2018). CSC referral could also facilitate multi-agency work, allowing previously unmet health needs to be identified and addressed. Multi-agency working is at the heart of the government’s reforms of Children’s Social Care. New legislation set out in the Children’s Wellbeing and Schools Bill, currently undergoing examination in the House of Lords, will require each local authority to set up a multi-agency child protection team (UK Parliament, 2025). In addition, new policy guidance issued in March 2025 by the DfE sets the expectation that all local areas put in place a multi-disciplinary Family Help team, with the vision of a single holistic assessment and plan for a child and family, which may include triggering or coordinating healthcare provision (Department for Education, 2025). The Family Help teams will work with families meeting Child in Need thresholds or eligible for Targeted Early Help (children and families with multiple or complex needs that require a plan to be in place and a lead practitioner). Like other recent research (Mendez Pineda et al., 2025), our findings suggest that there may already be current practice to understand and build on for implementing these reforms to improve outcomes and experiences of children and families. On the health policy side, the Department of Health and Social Care in England has made a very strong commitment to prevention (NHS, 2025), which is consistent with a move from unplanned to planned healthcare for children and families. A next step for research is to investigate variation across local areas and whether contact with CSC appears to affect the health care use of other family members, in keeping with the whole family approach of Family Help teams. If some local areas have stronger shifts from unplanned to planned healthcare use after contact with CSC, we may be able to examine and learn from what is ‘going right’ in terms of multi-agency working in these areas, to support other areas in their implementation of the Children’s Social Care reforms.

## 5. Conclusion

Taken together, our findings suggest that CSC involvement may uncover unmet healthcare needs and facilitate more structured healthcare engagement, particularly for planned care, potentially offering a window for early intervention and coordinated care responses to children’s health needs. This shift from unplanned to planned care could potentially prevent avoidable unplanned healthcare utilisation in the long term. However, it is unclear to what extent this shift reflects the effects of CSC involvement and multi-agency work or changes in children’s environment. Future work should examine the drivers of this shift, variation across local levels, investigate areas which appear to have strong shifts to planned care for children involved with CSC and include wider family members in the scope of enquiry.

## Data Availability

ECHILD is available for access through the Office for National Statistics Secure Research Service (ONS SRS) under the Digital Economy Act (DEA). This means that accredited researchers can apply to access the data for approved projects that serve the public good, in accordance with the legal safeguards and governance requirements set out by the DEA. Access is granted only to researchers who meet the accreditation criteria and agree to comply with all data protection, ethical standards and requirements set by data owners and data controllers.

https://www.echild.ac.uk/governance

## Funding

This work was supported by the Economic and Social Research Council (ESRC) through the ADR England Community Catalyst: Children at risk of poor outcomes (ES/Y010566/1).

## Declaration of interest

The authors declare that they have no conflicts of interest relevant to this work.

## Author contributions

EL: Conceptualization, data curation, formal analysis, investigation, methodology, visualizations, writing – original draft, review and editing; JW: Conceptualization, review and editing, supervision; KH: Conceptualization, review and editing, supervision, funding acquisition; LJG: Conceptualization, review and editing, funding acquisition; KB: Conceptualization, review and editing, funding acquisition; DH: Conceptualization, review and editing; LH: Conceptualization, review and editing; KT: Conceptualization, review and editing; GB: Conceptualization, review and editing

## Notes

### Competing Interest Statement

The authors have declared no competing interest.

### Author Declarations

The governance of the ECHILD Database is supported by a series of approvals and ethical oversight mechanisms that ensure the responsible use of linked administrative data for specific research purposes. Approvals to create and evaluate ECHILD were granted by the following bodies: Department for Education (DfE): DR200604.02 NHS England (NHSE): DARS-NIC-381972 Ethical approval for the ECHILD project was granted by: National Research Ethics Service: 17/LO/1494 NHS Health Research Authority Research Ethics Committee: 20/EE/0180 and 21/SW/0159 UCL Great Ormond Street Institute of Child Healths Joint Research and Development Office: 20PE16

