## Supplementary material for "Healthcare utilisation among children in contact with social services in England: an interrupted time series using ECHILD"

#### Supplementary Material A

Flow of sample selection for each CSC group, showing sequential exclusions and final analytic sample sizes for Children in Need (CiN), Child on Protection Plans (CPP), and Children Looked After (CLA).

##### Table S1: Process of sample selection by CSC group

| Steps | CiN N* | CPP N | CLA N |
|---|---|---|---|
| 1: Original sample | 11,633,180 | 11,633,180 | 3,181,460 |
| Exclusion due to missing referral date | 53,220 | 53,220 | 0 |
| 2: N of observations with non-missing referral date | 11,579,960 | 11,579,960 | 3,181,460 |
| Exclusion due to unlikely date of birth | 11,940 | 11,940 | 0 |
| 3: N of observations with valid date of birth | 11,568,020 | 11,568,020 | 3,181,460 |
| Exclusion of those aged < 2 or ≥ 18 years old | 1,798,940 | 1,798,940 | 494,570 |
| 4: Observation of children aged ≥ 2 and < 18 years old | 9,769,080 | 9,769,080 | 2,686,890 |
| Exclusion due to being referred to CSC before 01/04/2009 and after 31/03/2018 | 3,087,220 | 3,087,220 | 1,483,660 |
| 5: Observations of children referred to CSC between 01/04/2009 and 31/03/2018 | 6,681,860 | 6,681,860 | 1,203,230 |
| Exclusions due to having a different CSC status than the analysis group** | 613,440 | 6,068,410 | - |
| 6: Observations with CSC status matching the analysis group | 6,068,410 | 613,440 | - |
| Exclusions due to no CSC assessment | 591,430 | - | - |
| 7: Observations of children who were assessed by CSC | 5,476,980 | - | - |
| Exclusions of those assessed but not found in need | 1,631,670 | - | - |
| 8: Observations of children who were identified as CiN by CSC | 3,845,310 | - | - |
| Exclusion of observations that were not involved with CSC for the first time during period | 2,128,650 | 324,300 | 955,940 |
| 9: Children involved with CSC for the first time during the period | 1,716,660 | 289,140 | 247,290 |
| Exclusion of observations with missing aPMR | 311,960 | 27,880 | 48,250 |
| 10: Observations with non-missing aPMR | 1,404,700 | 261,270 | 199,040 |
| Exclusion of observations with duplicated aPMR | 92,070 | 12,160 | 8,400 |
| 11: Observations with unique aPMR | 1,312,620 | 249,110 | 190,630 |
| Exclusions of observations with missing age, ethnicity or deprivation | 49,310 | 970 | 760 |
| 12: Observation with non-missing age, ethnicity or deprivation | 1,263,310 | 248,140 | 189,880 |
| Exclusion of observations that had other CSC status within the examined period | 191,390 | 34,530 | - |
| 13: Observations with non-overlapping CSC statuses within the period | 1,071,920 | 213,610 | - |
| Exclusions due to non-linkage with HES data | 57,590 | 9,370 | 12,240 |
| 14: Final sample size | 1,014,330 | 204,240 | 177,640 |

**CiN and CPP are from the same dataset, which is why their numbers are identical up to step 5*

*** A different CSC status refers to a status inconsistent with the analysis group—for example, CPP status is considered different for the CiN group, and CiN status is considered different for the CPP group*

### Supplementary Material B

Linkage rates between CSC datasets and Hospital Episode Statistics (HES) by CSC group and financial year, showing consistently high linkage across the study period.

#### Table S2. Linkage rates by CSC groups and financial year

| Financial year | CiN | | | CPP | | | CLA* | | |
|---|---|---|---|---|---|---|---|---|---|
| | Total N | Linked to HES | Linkage rate (%) | Total N | Linked to HES | Linkage rate (%) | Total N | Linked to HES | Linkage rate (%) |
| 2009-2010 | 184,320 | 171,840 | 93.2 | 2,860 | 2,730 | 95.5 | 36,830 | 34,060 | 93 |
| 2010-2011 | 131,730 | 123,610 | 93.8 | 8,340 | 7,860 | 94.2 | 22,970 | 21,400 | 93 |
| 2011-2012 | 126,030 | 118,750 | 94.2 | 22,340 | 21,120 | 94.5 | 19,830 | 18,550 | 94 |
| 2012-2013 | 125,340 | 118,310 | 94.4 | 26,940 | 25,510 | 94.7 | 18,640 | 17,460 | 94 |
| 2013-2014 | 120,650 | 114,620 | 95.0 | 30,000 | 28,710 | 95.7 | 18,890 | 17,770 | 94 |
| 2014-2015 | 100,300 | 95,530 | 95.2 | 29,760 | 28,480 | 95.7 | 18,620 | 17,460 | 94 |
| 2015-2016 | 95,000 | 90,700 | 95.5 | 29,500 | 28,290 | 95.9 | 17,980 | 16,840 | 94 |
| 2016-2017 | 93,030 | 89,170 | 95.9 | 30,750 | 29,600 | 96.3 | 18,400 | 17,320 | 94 |
| 2017-2018 | 95,530 | 91,910 | 96.2 | 33,110 | 31,950 | 96.5 | 17,720 | 16,800 | 95 |

**CLA percentages has been rounded to 0 d.p in compliance with DfE statistical disclosure policy*

### Supplementary Material C

Baseline demographic, socioeconomic, and referral characteristics of children in each CSC group prior to HES linkage, including age, sex, ethnicity, primary need, referral source, and deprivation.

#### Table S3a. Comparison of sociodemographic characteristics of Children in Need before and after linkage to HES

| Variable | Category | Unlinked sample | | Linked sample | |
|---|---|---|---|---|---|
| | | CiN (N = 1,071,920) | | CiN (N = 1,014,330) | |
| | | N | % | N | % |
| **Continuous age** | Age (mean years, SD) | 9.7 | (4.4) | 9.7 | (4.4) |
| **Age group** | 2-4 years old | 190,680 | 17.8 | 182,590 | 18.0 |
| | 5-11 years old | 508,300 | 47.4 | 485,060 | 47.8 |
| | 12-17 years old | 372,940 | 34.8 | 346,680 | 34.2 |
| **Ethnicity** | Any other ethnic group | 21,440 | 2.0 | 20,130 | 2.0 |
| | Asian | 89,080 | 8.3 | 83,580 | 8.2 |
| | Black | 81,500 | 7.6 | 73,840 | 7.3 |
| | Mixed | 64,940 | 6.1 | 60,320 | 5.9 |
| | White | 814,960 | 76 | 776,460 | 76.5 |
| **Sex** | Male | 545,020 | 50.8 | 520,130 | 51.3 |
| | Female | 526,900 | 49.2 | 494,210 | 48.7 |
| **Primary need** | Abuse or neglect | 547,330 | 51.1 | 519,510 | 51.2 |

| | | | | | |
|---|---|---|---|---|---|
| | Child's disability/illness | 64,320 | 6.0 | 60,990 | 6.0 |
| | Parental disability/illness | 24,670 | 2.3 | 23,280 | 2.3 |
| | Family in acute stress | 113,310 | 10.6 | 106,540 | 10.5 |
| | Family dysfunction | 199,230 | 18.6 | 188,440 | 18.6 |
| | Socially unacceptable | 29,300 | 2.7 | 27,690 | 2.7 |
| | Low income/ Absent parenting | 14,470 | 1.3 | 13,220 | 1.3 |
| | Cases other than CiN | 14,510 | 1.4 | 13,460 | 1.3 |
| | NA | 64,780 | 6.0 | 61,210 | 6.0 |
| **Source** | Individual | 57,040 | 5.3 | 54,120 | 5.3 |
| | Schools | 132,680 | 12.4 | 126,850 | 12.5 |
| | Health services | 66,740 | 6.2 | 63,640 | 6.3 |
| | Housing | 6,380 | 0.6 | 6,070 | 0.6 |
| | LA services | 63,180 | 5.9 | 60,090 | 5.9 |
| | Police | 140,800 | 13.1 | 134,660 | 13.3 |
| | Other legal agency | 14,920 | 1.4 | 14,220 | 1.4 |
| | Other | 34,140 | 3.2 | 32,420 | 3.2 |
| | Anonymous | 9,240 | 0.9 | 8,960 | 0.9 |
| | NA | 546,790 | 51.0 | 513,300 | 50.6 |
| IMD Decile | Most deprived | 135,620 | 12.7 | 127,410 | 12.6 |
| | 2 | 89,000 | 8.3 | 83,490 | 8.2 |

| | | | | | |
|---|---|---|---|---|---|
| | 3 | 87,000 | 8.1 | 81,890 | 8.1 |
| | 4 | 101,480 | 9.5 | 96,770 | 9.5 |
| | 5 | 96,930 | 9.0 | 92,400 | 9.1 |
| | 6 | 112,400 | 10.5 | 106,390 | 10.5 |
| | 7 | 157,740 | 14.7 | 149,930 | 14.8 |
| | 8 | 100,900 | 9.4 | 94,780 | 9.3 |
| | 9 | 115,560 | 10.8 | 109,620 | 10.8 |
| | Least deprived | 75,300 | 7.0 | 71,670 | 7.1 |

#### Table S3b. Comparison of sociodemographic characteristics of Children on Protection Plans before and after linkage to HES

| Variable | Category | Unlinked sample | | Linked sample | |
|---|---|---|---|---|---|
| | | CPP (N = 213,610) | | CPP (N = 204,240) | |
| | | **N** | **%** | **N** | **%** |
| **Continuous age** | Age (mean years, SD) | 8.4 | (4.0) | 8.4 | (4.0) |
| **Age group** | 2-4 years old | 53,930 | 25.2 | 51,960 | 25.4 |
| | 5-11 years old | 110,670 | 51.8 | 105,920 | 51.9 |
| | 12-17 years old | 49,010 | 22.9 | 46,360 | 22.7 |
| **Ethnicity** | Any other ethnic group | 3,080 | 1.4 | 2,940 | 1.4 |
| | Asian | 15,250 | 7.1 | 14,500 | 7.1 |
| | Black | 12,170 | 5.7 | 11,230 | 5.5 |
| | Mixed | 17,160 | 8.0 | 16,250 | 8 |
| | White | 165,960 | 77.7 | 159,320 | 78 |
| **Sex** | Male | 108,550 | 50.8 | 104,130 | 51 |
| | Female | 105,060 | 49.2 | 100,100 | 49 |
| **Primary need** | Abuse or neglect | 132,170 | 61.9 | 126,480 | 61.9 |
| | Child's disability/illness | 3,730 | 1.7 | 3,550 | 1.7 |
| | Parental disability/illness | 5,830 | 2.7 | 5,550 | 2.7 |
| | Family in acute stress | 18,030 | 8.4 | 17,130 | 8.4 |

| | | | | | |
|---|---|---|---|---|---|
| | Family dysfunction | 41,660 | 19.5 | 39,860 | 19.5 |
| | Socially unacceptable | 3,650 | 1.7 | 3,500 | 1.7 |
| | Low income/ Absent parenting | 1,350 | 0.6 | 1,290 | 0.6 |
| | Cases other than CiN | 1,330 | 0.6 | 1,270 | 0.6 |
| | NA | 5,860 | 2.7 | 5,600 | 2.7 |
| **Source** | Individual | 13,140 | 6.2 | 12,540 | 6.1 |
| | Schools | 39,350 | 18.4 | 37,740 | 18.5 |
| | Health services | 20,690 | 9.7 | 19,800 | 9.7 |
| | Housing | 1,940 | 0.9 | 1,860 | 0.9 |
| | LA services | 28,720 | 13.4 | 27,420 | 13.4 |
| | Police | 48,860 | 22.9 | 46,820 | 22.9 |
| | Other legal agency | 4,670 | 2.2 | 4,480 | 2.2 |
| | Other | 11,580 | 5.4 | 11,100 | 5.4 |
| | Anonymous | 4,070 | 1.9 | 3,900 | 1.9 |
| | NA | 40,590 | 19.0 | 38,580 | 18.9 |
| IMD Decile | Most deprived | 27,830 | 13.0 | 26,460 | 13.0 |
| | 2 | 19,200 | 9.0 | 18,260 | 8.9 |
| | 3 | 15,990 | 7.5 | 15,180 | 7.4 |
| | 4 | 21,040 | 9.9 | 20,220 | 9.9 |
| | 5 | 20,210 | 9.5 | 19,460 | 9.5 |

| | | | | | |
|---|---|---|---|---|---|
| | 6 | 18,050 | 8.5 | 17,240 | 8.4 |
| | 7 | 31,650 | 14.8 | 30,420 | 14.9 |
| | 8 | 19,450 | 9.1 | 18,560 | 9.1 |
| | 9 | 24,220 | 11.3 | 23,220 | 11.4 |
| | Least deprived | 15,960 | 7.5 | 15,230 | 7.5 |

#### Table S3c. Comparison of sociodemographic characteristics of Children Looked After before and after linkage to HES

| Variable | Category | Unlinked sample | | Linked sample | |
|---|---|---|---|---|---|
| | | CLA* (N = 189,880) | | CLA (N = 177,640) | |
| | | **N** | **%** | **N** | **%** |
| **Continuous age** | Age (mean years, SD) | 10.9 | (4.7) | 10.9 | (4.7) |
| **Age group** | 2-4 years old | 30,360 | 16 | 28,560 | 16 |
| | 5-11 years old | 67,160 | 35 | 63,920 | 36 |
| | 12-17 years old | 92,360 | 49 | 85,150 | 48 |
| **Ethnicity** | Any other ethnic group | 4,290 | 2 | 3,900 | 2 |
| | Asian | 10,860 | 6 | 10,020 | 6 |
| | Black | 16,760 | 9 | 15,090 | 8 |
| | Mixed | 16,250 | 9 | 15,010 | 8 |
| | White | 141,720 | 75 | 133,620 | 75 |
| **Sex** | Male | 102,010 | 54 | 96,320 | 54 |
| | Female | 87,870 | 46 | 81,320 | 46 |
| **Primary need** | Abuse or neglect | 104,830 | 55 | 98,300 | 55 |
| | Child's disability/illness | 11,140 | 6 | 10,640 | 6 |
| | Parental disability/illness | 6,050 | 3 | 5,680 | 3 |
| | Family in acute stress | 19,860 | 10 | 18,410 | 10 |

| | | | | | |
|---|---|---|---|---|---|
| | Family dysfunction | 31,800 | 17 | 29,710 | 17 |
| | Socially unacceptable | 6,920 | 4 | 6,470 | 4 |
| | Low income/ Absent parenting | 9,280 | 5 | 8,440 | 5 |
| | Cases other than CiN | - | - | - | - |
| | NA | - | - | - | - |
| IMD Decile | Most deprived | 26,770 | 14 | 25,080 | 14 |
| | 2 | 17,040 | 9 | 15,790 | 9 |
| | 3 | 16,730 | 9 | 15,580 | 9 |
| | 4 | 19,580 | 10 | 18,400 | 10 |
| | 5 | 18,630 | 10 | 17,550 | 10 |
| | 6 | 17,960 | 9 | 16,780 | 9 |
| | 7 | 24,000 | 13 | 22,600 | 13 |
| | 8 | 18,920 | 10 | 17,540 | 10 |
| | 9 | 19,440 | 10 | 18,230 | 10 |
| | Least deprived | 10,800 | 6 | 10,090 | 6 |

** CLA percentages have been rounded to 0 d.p in compliance with DfE statistical disclosure policy*

### Supplementary Material D

Hospital contacts by CSC group and matched peers: number of hospital contacts by period by care type before and after referral, plus rate ratios comparing CSC groups with matched cohorts across periods.

#### Table S4: Number of hospital contacts by period, group and health service

| Type of care | Health Service | Group | Number of hospital contacts 2 years pre CSC referral | | Number of hospital contacts 2 years post CSC referral | |
|---|---|---|---|---|---|---|
| | | | Children involved with CSC | Matched cohort | Children involved with CSC | Matched cohort |
| Planned care | A&E Attendances | CiN | 920,420 | 579,100 | 967,260 | 568,420 |
| | | CPP | 196,650 | 131,200 | 199,660 | 121,400 |
| | | CLA | 200,720 | 102,660 | 253,210 | 101,200 |
| | Unplanned admissions | CiN | 203,380 | 92,540 | 194,010 | 75,070 |
| | | CPP | 38,980 | 22,120 | 38,120 | 16,400 |
| | | CLA | 55,240 | 16,050 | 54,500 | 13,420 |
| Unplanned care | Planned admissions | CiN | 176,200 | 82,770 | 221,460 | 86,370 |
| | | CPP | 23,990 | 17,420 | 31,080 | 17,520 |
| | | CLA | 39,000 | 14,010 | 42,900 | 15,430 |
| | Outpatient appointments | CiN | 2,846,830 | 1,534,570 | 3,439,970 | 1,678,940 |
| | | CPP | 498,250 | 324,670 | 690,390 | 350,290 |
| | | CLA | 630,120 | 275,480 | 827,180 | 304,620 |
| Combined | Total hospital contacts matched | CiN | 4,146,830 | 2,288,990 | 4,822,700 | 2,408,790 |
| | | CPP | 757,860 | 495,410 | 959,250 | 505,600 |
| | | CLA | 925,080 | 408,190 | 1,177,790 | 434,670 |

#### Table S5: Rate ratio of monthly rate of hospital contacts by group

| Comparison | Health Service | Rate Ratio of period between months -24 to -3 | Rate ratio of period between months -2 to 2 | Rate ratio of period between months 3 to 23 |
|---|---|---|---|---|
| **Children in Need compared to their matched group** | A&E | 1.54 | 1.98 | 1.67 |
| | Unplanned admissions | 2.06 | 3.64 | 2.45 |
| | Planned admissions | 2.08 | 2.87 | 2.50 |
| | Outpatient | 1.83 | 2.24 | 2.01 |
| | **Combined** | **1.88** | **2.68** | **2.16** |
| **Children on Protection Plans compared to their matched group** | A&E | 1.47 | 1.84 | 1.61 |
| | Unplanned admissions | 1.70 | 2.96 | 2.18 |
| | Planned admissions | 1.36 | 1.64 | 1.79 |
| | Outpatient | 1.53 | 1.89 | 1.96 |
| | **Combined** | **1.52** | **2.08** | **1.89** |
| **Children Looked After compared to their matched group** | A&E | 1.84 | 3.08 | 2.43 |
| | Unplanned admissions | 3.06 | 6.82 | 3.79 |
| | Planned admissions | 2.75 | 2.95 | 2.77 |
| | Outpatient | 2.24 | 3.44 | 2.56 |
| | **Combined** | **2.47** | **4.07** | **2.89** |

### Supplementary Material E

Negative binomial regression estimates for monthly rates of hospital contacts across CSC groups (CiN, CPP, CLA), presented separately by care type (A&E attendances, unplanned admissions, planned admissions, outpatient appointments), with and without covariate adjustment.

#### Table S6. Negative binomial regression estimates for CiN (n = 1,014,330) A&E attendances

| Variable | Model 1: No covariates | | | | Model 2: Adjusted by covariates | | | |
|---|---|---|---|---|---|---|---|---|
| | Coefficient | p-value | Lower 95% CI | Upper 95% CI | Coefficient | p-value | Lower 95% CI | Upper 95% CI |
| **(Intercept)** | 0.039 | <0.001 | 0.038 | 0.040 | 0.064 | < 0.001 | 0.062 | 0.065 |
| **Month** | 1.005 | <0.001 | 1.003 | 1.006 | 1.003 | < 0.001 | 1.001 | 1.004 |
| **Intervention** | 1.003 | 0.807 | 0.980 | 1.027 | 0.991 | 0.571 | 0.962 | 1.021 |
| **Month x Intervention** | 0.995 | <0.001 | 0.993 | 0.998 | 0.996 | < 0.001 | 0.994 | 0.998 |
| **Age group (ref: 2 – 4.99) 5 - 11.99** | - | - | - | - | 0.615 | < 0.001 | 0.606 | 0.625 |
| **Age group 12 - 17.99** | - | - | - | - | 0.818 | < 0.001 | 0.802 | 0.835 |
| **IMD decile** | - | - | - | - | 0.975 | < 0.001 | 0.973 | 0.977 |
| **Female** | - | - | - | - | 0.907 | < 0.001 | 0.894 | 0.919 |
| **Ethnicity (ref: White) Any other ethnic group** | - | - | - | - | 0.970 | 0.006 | 0.950 | 0.991 |
| **Asian** | - | - | - | - | 0.879 | < 0.001 | 0.861 | 0.896 |
| **Black** | - | - | - | - | 0.811 | < 0.001 | 0.795 | 0.827 |
| **Mixed** | - | - | - | - | 0.936 | < 0.001 | 0.918 | 0.955 |

#### Table S7. Negative binomial regression estimates for CPP (n = 204,240) A&E attendances

| Variable | Model 1: No covariates | | | | Model 2: Adjusted by covariates | | | |
|---|---|---|---|---|---|---|---|---|
| | Coefficient | p-value | Lower 95% CI | Upper 95% CI | Coefficient | p-value | Lower 95% CI | Upper 95% CI |
| **(Intercept)** | 0.039 | < 0.001 | 0.039 | 0.040 | 0.058 | < 0.001 | 0.056 | 0.059 |
| **Month** | 0.999 | 0.018 | 0.998 | 1.000 | 0.998 | 0.054 | 0.997 | 1.000 |
| **Intervention** | 1.081 | < 0.001 | 1.049 | 1.114 | 1.092 | < 0.001 | 1.049 | 1.136 |
| **Month x Intervention** | 0.996 | 0.005 | 0.994 | 0.999 | 0.996 | 0.011 | 0.993 | 0.999 |
| **Age group (ref: 2 – 4.99) 5 - 11.99** | - | - | - | - | 0.651 | < 0.001 | 0.639 | 0.664 |
| **Age group 12 - 17.99** | - | - | - | - | 0.955 | < 0.001 | 0.929 | 0.982 |
| **IMD decile** | - | - | - | - | 0.978 | < 0.001 | 0.975 | 0.980 |
| **Female** | - | - | - | - | 0.935 | < 0.001 | 0.918 | 0.952 |
| **Ethnicity (ref: White) Any other ethnic group** | - | - | - | - | 1.035 | 0.059 | 0.999 | 1.072 |
| **Asian** | - | - | - | - | 0.935 | < 0.001 | 0.913 | 0.958 |
| **Black** | - | - | - | - | 0.844 | < 0.001 | 0.822 | 0.866 |
| **Mixed** | - | - | - | - | 0.936 | < 0.001 | 0.914 | 0.957 |

#### Table S8. Negative binomial regression estimates for CLA (n = 177,640) A&E attendances

| Variable | Model 1: No covariates | | | | Model 2: Adjusted by covariates | | | |
|---|---|---|---|---|---|---|---|---|
| | Coefficient | p-value | Lower 95% CI | Upper 95% CI | Coefficient | p-value | Lower 95% CI | Upper 95% CI |
| (Intercept) | 0.057 | <0.001 | 0.054 | 0.060 | 0.047 | <0.001 | 0.045 | 0.050 |
| Month | 1.019 | <0.001 | 1.016 | 1.023 | 1.012 | <0.001 | 1.009 | 1.014 |
| Intervention | 1.039 | 0.138 | 0.988 | 1.092 | 1.002 | 0.955 | 0.945 | 1.061 |
| Month x Intervention | 0.979 | <0.001 | 0.974 | 0.984 | 0.985 | <0.001 | 0.981 | 0.989 |
| Age group (ref: 2 – 4.99) 5 - 11.99 | - | - | - | - | 0.859 | <0.001 | 0.830 | 0.889 |
| Age group 12 - 17.99 | - | - | - | - | 1.826 | <0.001 | 1.758 | 1.898 |
| IMD decile | - | - | - | - | 0.968 | <0.001 | 0.964 | 0.972 |
| Female | - | - | - | - | 1.080 | <0.001 | 1.052 | 1.108 |
| Ethnicity (ref: White) Any other ethnic group | - | - | - | - | 0.661 | <0.001 | 0.626 | 0.698 |
| Asian | - | - | - | - | 0.799 | <0.001 | 0.768 | 0.832 |
| Black | - | - | - | - | 0.781 | <0.001 | 0.755 | 0.809 |
| Mixed | - | - | - | - | 0.917 | <0.001 | 0.887 | 0.947 |

#### Table S9. Negative binomial regression estimates for CiN (n = 1,014,330) unplanned admissions

| Variable | Model 1: No covariates | | | | Model 2: Adjusted by covariates | | | |
|---|---|---|---|---|---|---|---|---|
| | Coefficient | p-value | Lower 95% CI | Upper 95% CI | Coefficient | p-value | Lower 95% CI | Upper 95% CI |
| **(Intercept)** | 0.008 | < 0.001 | 0.007 | 0.009 | 0.014 | < 0.001 | 0.014 | 0.015 |
| **Month** | 0.999 | 0.683 | 0.993 | 1.005 | 0.999 | 0.715 | 0.997 | 1.002 |
| **Intervention** | 1.060 | 0.161 | 0.977 | 1.151 | 1.028 | 0.330 | 0.973 | 1.086 |
| **Month x Intervention** | 0.994 | 0.075 | 0.986 | 1.001 | 0.994 | 0.003 | 0.989 | 0.998 |
| **Age group (ref: 2 – 4.99) 5 - 11.99** | - | - | - | - | 0.415 | < 0.001 | 0.405 | 0.425 |
| **Age group 12 - 17.99** | - | - | - | - | 0.597 | < 0.001 | 0.576 | 0.618 |
| **IMD decile** | - | - | - | - | 0.994 | < 0.001 | 0.990 | 0.997 |
| **Female** | - | - | - | - | 1.011 | 0.386 | 0.987 | 1.035 |
| **Ethnicity (ref: White) Any other ethnic group** | - | - | - | - | 0.836 | < 0.001 | 0.803 | 0.870 |
| **Asian** | - | - | - | - | 1.051 | 0.002 | 1.019 | 1.085 |
| **Black** | - | - | - | - | 0.736 | < 0.001 | 0.712 | 0.760 |
| **Mixed** | - | - | - | - | 0.852 | < 0.001 | 0.824 | 0.881 |

#### Table S10. Negative binomial regression estimates for CPP (n = 204,240) unplanned admissions

| Variable | Model 1: No covariates | | | | Model 2: Adjusted by covariates | | | |
|---|---|---|---|---|---|---|---|---|
| | Coefficient | p-value | Lower 95% CI | Upper 95% CI | Coefficient | p-value | Lower 95% CI | Upper 95% CI |
| (Intercept) | 0.007 | <0.001 | 0.006 | 0.007 | 0.011 | < 0.001 | 0.010 | 0.012 |
| Month | 0.989 | <0.001 | 0.985 | 0.992 | 0.991 | < 0.001 | 0.987 | 0.995 |
| Intervention | 1.307 | <0.001 | 1.223 | 1.397 | 1.298 | < 0.001 | 1.196 | 1.409 |
| Month x Intervention | 0.996 | 0.308 | 0.990 | 1.003 | 0.995 | 0.092 | 0.988 | 1.001 |
| Age group (ref: 2 – 4.99) 5 - 11.99 | - | - | - | - | 0.443 | < 0.001 | 0.427 | 0.459 |
| Age group 12 - 17.99 | - | - | - | - | 0.782 | < 0.001 | 0.741 | 0.826 |
| IMD decile | - | - | - | - | 0.988 | < 0.001 | 0.983 | 0.994 |
| Female | - | - | - | - | 1.033 | 0.076 | 0.997 | 1.072 |
| Ethnicity (ref: White) Any other ethnic group | - | - | - | - | 0.956 | 0.273 | 0.883 | 1.036 |
| Asian | - | - | - | - | 1.032 | 0.175 | 0.986 | 1.080 |
| Black | - | - | - | - | 0.783 | < 0.001 | 0.743 | 0.825 |
| Mixed | - | - | - | - | 0.899 | < 0.001 | 0.859 | 0.940 |

#### Table S11. Negative binomial regression estimates for CLA (n = 177,640) unplanned admissions

| Variable | Model 1: No covariates | | | | Model 2: Adjusted by covariates | | | |
|---|---|---|---|---|---|---|---|---|
| | Coefficient | p-value | Lower 95% CI | Upper 95% CI | Coefficient | p-value | Lower 95% CI | Upper 95% CI |
| **(Intercept)** | 0.015 | < 0.001 | 0.013 | 0.017 | 0.013 | < 0.001 | 0.012 | 0.014 |
| **Month** | 1.020 | < 0.001 | 1.011 | 1.028 | 1.011 | < 0.001 | 1.006 | 1.015 |
| **Intervention** | 0.890 | 0.034 | 0.799 | 0.992 | 0.903 | 0.030 | 0.824 | 0.990 |
| **Month x Intervention** | 0.971 | < 0.001 | 0.962 | 0.981 | 0.978 | < 0.001 | 0.971 | 0.985 |
| **Age group (ref: 2 – 4.99) 5 - 11.99** | - | - | - | - | 0.618 | < 0.001 | 0.586 | 0.653 |
| **Age group 12 - 17.99** | - | - | - | - | 1.340 | < 0.001 | 1.266 | 1.418 |
| **IMD decile** | - | - | - | - | 0.988 | < 0.001 | 0.982 | 0.994 |
| **Female** | - | - | - | - | 1.340 | < 0.001 | 1.288 | 1.393 |
| **Ethnicity (ref: White) Any other ethnic group** | - | - | - | - | 0.556 | < 0.001 | 0.508 | 0.609 |
| **Asian** | - | - | - | - | 0.860 | < 0.001 | 0.810 | 0.913 |
| **Black** | - | - | - | - | 0.652 | < 0.001 | 0.619 | 0.686 |
| **Mixed** | - | - | - | - | 0.815 | < 0.001 | 0.775 | 0.856 |

#### Table S12. Negative binomial regression estimates for CiN (n = 1,014,330) planned admissions

| Variable | Model 1: No covariates | | | | Model 2: Adjusted by covariates | | | |
|---|---|---|---|---|---|---|---|---|
| | Coefficient | p-value | Lower 95% CI | Upper 95% CI | Coefficient | p-value | Lower 95% CI | Upper 95% CI |
| **(Intercept)** | 0.008 | < 0.001 | 0.007 | 0.008 | 0.010 | <0.001 | 0.009 | 0.010 |
| **Month** | 1.007 | 0.001 | 1.003 | 1.012 | 1.007 | <0.001 | 1.005 | 1.010 |
| **Intervention** | 1.260 | < 0.001 | 1.182 | 1.343 | 1.246 | <0.001 | 1.184 | 1.311 |
| **Month x Intervention** | 0.986 | < 0.001 | 0.980 | 0.992 | 0.984 | <0.001 | 0.981 | 0.988 |
| **Age group (ref: 2 – 4.99) 5 - 11.99** | - | - | - | - | 0.697 | <0.001 | 0.681 | 0.714 |
| **Age group 12 - 17.99** | - | - | - | - | 0.728 | <0.001 | 0.706 | 0.751 |
| **IMD decile** | - | - | - | - | 1.010 | <0.001 | 1.007 | 1.014 |
| **Female** | - | - | - | - | 1.014 | 0.218 | 0.992 | 1.036 |
| **Ethnicity (ref: White) Any other ethnic group** | - | - | - | - | 1.014 | 0.514 | 0.973 | 1.056 |
| **Asian** | - | - | - | - | 1.258 | <0.001 | 1.222 | 1.294 |
| **Black** | - | - | - | - | 0.923 | <0.001 | 0.894 | 0.953 |
| **Mixed** | - | - | - | - | 0.813 | <0.001 | 0.785 | 0.842 |

#### Table S13. Negative binomial regression estimates for CPP (n = 204,240) planned admissions

| Variable | Model 1: No covariates | | | | Model 2: Adjusted by covariates | | | |
|---|---|---|---|---|---|---|---|---|
| | Coefficient | p-value | Lower 95% CI | Upper 95% CI | Coefficient | p-value | Lower 95% CI | Upper 95% CI |
| **(Intercept)** | 0.005 | <0.001 | 0.004 | 0.005 | 0.005 | < 0.001 | 0.005 | 0.006 |
| **Month** | 0.997 | 0.445 | 0.989 | 1.005 | 0.996 | 0.063 | 0.991 | 1.000 |
| **Intervention** | 1.339 | <0.001 | 1.193 | 1.504 | 1.410 | < 0.001 | 1.295 | 1.536 |
| **Month x Intervention** | 1.005 | 0.222 | 0.997 | 1.013 | 1.007 | 0.034 | 1.001 | 1.013 |
| **Age group (ref: 2 – 4.99) 5 - 11.99** | - | - | - | - | 0.854 | < 0.001 | 0.819 | 0.891 |
| **Age group 12 - 17.99** | - | - | - | - | 0.945 | 0.028 | 0.899 | 0.994 |
| **IMD decile** | - | - | - | - | 0.987 | < 0.001 | 0.981 | 0.993 |
| **Female** | - | - | - | - | 0.917 | < 0.001 | 0.885 | 0.951 |
| **Ethnicity (ref: White) Any other ethnic group** | - | - | - | - | 0.987 | 0.766 | 0.909 | 1.073 |
| **Asian** | - | - | - | - | 1.240 | < 0.001 | 1.175 | 1.309 |
| **Black** | - | - | - | - | 0.959 | 0.114 | 0.910 | 1.010 |
| **Mixed** | - | - | - | - | 1.047 | 0.153 | 0.983 | 1.116 |

#### Table S14. Negative binomial regression estimates for CLA (n = 177,640) planned admissions

| Variable | Model 1: No covariates | | | | Model 2: Adjusted by covariates | | | |
|---|---|---|---|---|---|---|---|---|
| | Coefficient | p-value | Lower 95% CI | Upper 95% CI | Coefficient | p-value | Lower 95% CI | Upper 95% CI |
| **(Intercept)** | 0.010 | < 0.001 | 0.009 | 0.010 | 0.007 | < 0.001 | 0.007 | 0.008 |
| **Month** | 1.005 | 0.020 | 1.001 | 1.010 | 1.003 | 0.118 | 0.999 | 1.008 |
| **Intervention** | 1.115 | 0.042 | 1.004 | 1.237 | 1.234 | < 0.001 | 1.130 | 1.347 |
| **Month x Intervention** | 0.990 | < 0.001 | 0.986 | 0.995 | 0.991 | 0.006 | 0.985 | 0.997 |
| **Age group (ref: 2 – 4.99) 5 - 11.99** | - | - | - | - | 1.230 | < 0.001 | 1.168 | 1.295 |
| **Age group 12 - 17.99** | - | - | - | - | 1.319 | < 0.001 | 1.251 | 1.392 |
| **IMD decile** | - | - | - | - | 0.994 | 0.062 | 0.987 | 1.000 |
| **Female** | - | - | - | - | 1.152 | < 0.001 | 1.107 | 1.199 |
| **Ethnicity (ref: White) Any other ethnic group** | - | - | - | - | 0.557 | < 0.001 | 0.504 | 0.616 |
| **Asian** | - | - | - | - | 0.898 | < 0.001 | 0.844 | 0.955 |
| **Black** | - | - | - | - | 0.659 | < 0.001 | 0.622 | 0.698 |
| **Mixed** | - | - | - | - | 0.704 | < 0.001 | 0.664 | 0.746 |

#### Table S15. Negative binomial regression estimates for CiN (n = 1,014,330) outpatient appointments

| Variable | Model 1: No covariates | | | | Model 2: Adjusted by covariates | | | |
|---|---|---|---|---|---|---|---|---|
| | Coefficient | p-value | Lower 95% CI | Upper 95% CI | Coefficient | p-value | Lower 95% CI | Upper 95% CI |
| **(Intercept)** | 0.130 | < 0.001 | 0.127 | 0.132 | 0.140 | < 0.001 | 0.137 | 0.143 |
| **Month** | 1.009 | < 0.001 | 1.008 | 1.010 | 1.009 | < 0.001 | 1.008 | 1.010 |
| **Intervention** | 1.127 | < 0.001 | 1.103 | 1.152 | 1.178 | < 0.001 | 1.150 | 1.207 |
| **Month x Intervention** | 0.987 | < 0.001 | 0.985 | 0.990 | 0.986 | < 0.001 | 0.984 | 0.988 |
| **Age group (ref: 2 – 4.99) 5 - 11.99** | - | - | - | - | 0.715 | < 0.001 | 0.708 | 0.723 |
| **Age group 12 - 17.99** | - | - | - | - | 0.849 | < 0.001 | 0.837 | 0.861 |
| **IMD decile** | - | - | - | - | 1.035 | < 0.001 | 1.033 | 1.037 |
| **Female** | - | - | - | - | 0.895 | < 0.001 | 0.885 | 0.904 |
| **Ethnicity (ref: White) Any other ethnic group** | - | - | - | - | 0.970 | < 0.001 | 0.952 | 0.988 |
| **Asian** | - | - | - | - | 1.094 | < 0.001 | 1.078 | 1.109 |
| **Black** | - | - | - | - | 0.811 | < 0.001 | 0.800 | 0.823 |
| **Mixed** | - | - | - | - | 0.887 | < 0.001 | 0.873 | 0.901 |

#### Table S16. Negative binomial regression estimates for CPP (n = 204,240) outpatient appointments

| Variable | Model 1: No covariates | | | | Model 2: Adjusted by covariates | | | |
|---|---|---|---|---|---|---|---|---|
| | Coefficient | p-value | Lower 95% CI | Upper 95% CI | Coefficient | p-value | Lower 95% CI | Upper 95% CI |
| **(Intercept)** | 0.107 | < 0.001 | 0.106 | 0.109 | 0.107 | < 0.001 | 0.104 | 0.109 |
| **Month** | 1.005 | < 0.001 | 1.004 | 1.006 | 1.006 | < 0.001 | 1.005 | 1.007 |
| **Intervention** | 1.418 | < 0.001 | 1.374 | 1.464 | 1.482 | < 0.001 | 1.435 | 1.529 |
| **Month x Intervention** | 0.990 | < 0.001 | 0.988 | 0.991 | 0.987 | < 0.001 | 0.985 | 0.989 |
| **Age group (ref: 2 – 4.99) 5 - 11.99** | - | - | - | - | 0.832 | < 0.001 | 0.820 | 0.845 |
| **Age group 12 - 17.99** | - | - | - | - | 1.052 | < 0.001 | 1.033 | 1.072 |
| **IMD decile** | - | - | - | - | 1.032 | < 0.001 | 1.030 | 1.035 |
| **Female** | - | - | - | - | 0.865 | < 0.001 | 0.854 | 0.877 |
| **Ethnicity (ref: White) Any other ethnic group** | - | - | - | - | 0.971 | 0.061 | 0.941 | 1.001 |
| **Asian** | - | - | - | - | 1.025 | 0.009 | 1.006 | 1.045 |
| **Black** | - | - | - | - | 0.840 | < 0.001 | 0.824 | 0.856 |
| **Mixed** | - | - | - | - | 0.881 | < 0.001 | 0.867 | 0.896 |

#### Table S17. Negative binomial regression estimates for CLA (n = 177,640) outpatient appointments

| Variable | Model 1: No covariates | | | | Model 2: Adjusted by covariates | | | |
|---|---|---|---|---|---|---|---|---|
| | Coefficient | p-value | Lower 95% CI | Upper 95% CI | Coefficient | p-value | Lower 95% CI | Upper 95% CI |
| (Intercept) | 0.176 | < 0.001 | 0.169 | 0.182 | 0.148 | < 0.001 | 0.143 | 0.154 |
| Month | 1.015 | < 0.001 | 1.013 | 1.018 | 1.014 | < 0.001 | 1.012 | 1.016 |
| Intervention | 1.220 | < 0.001 | 1.145 | 1.301 | 1.332 | < 0.001 | 1.278 | 1.388 |
| Month x Intervention | 0.973 | < 0.001 | 0.968 | 0.979 | 0.973 | < 0.001 | 0.970 | 0.975 |
| Age group (ref: 2 – 4.99) 5 - 11.99 | - | - | - | - | 0.947 | < 0.001 | 0.927 | 0.968 |
| Age group 12 - 17.99 | - | - | - | - | 1.094 | < 0.001 | 1.070 | 1.120 |
| IMD decile | - | - | - | - | 1.025 | < 0.001 | 1.022 | 1.028 |
| Female | - | - | - | - | 1.026 | 0.004 | 1.009 | 1.044 |
| Ethnicity (ref: White) Any other ethnic group | - | - | - | - | 0.776 | < 0.001 | 0.748 | 0.805 |
| Asian | - | - | - | - | 0.920 | < 0.001 | 0.896 | 0.945 |
| Black | - | - | - | - | 0.774 | < 0.001 | 0.756 | 0.793 |
| Mixed | - | - | - | - | 0.854 | < 0.001 | 0.836 | 0.872 |
